# Person-centered measures of HCBS: Identifying existing approaches to measuring outcomes, services, and supports

**DOI:** 10.1101/2022.12.22.22283875

**Authors:** Lindsay DuBois, Niveda Tennety, Scott Roiter, Tonie Sadler, Dhrumil Shah, Sara Karon, Jessica Pedersen, Antoinette Nelson-Rodriguez, Q. Eileen Wafford, Allen Heinemann

## Abstract

This review aimed to describe the extent to which home- and community-based service (HCBS) outcome instruments published in the peer-reviewed literature are person-centered. We sought to identify measures that relate to progress toward or achievement of outcomes and service effectiveness. The research questions we evaluated were: (1) What nonmedical, person-centered outcome measures are used in HCBS? (2) What non-medical, person-centered effectiveness measures of HCBS supports are used? We worked with a research librarian to develop a bibliographic search strategy and identified text words and controlled vocabulary terms describing person-centered measures, non-medical context, home and community-based services, HCBS instruments, as well as effectiveness and quality. We performed the search on Medline (Ovid); The Cochrane Library (Wiley); CINAHL Plus with Full Text (Ebsco); Scopus (Elsevier); PsycInfo (Ebsco); and ProQuest dissertations & theses global. We searched each database from date of inception to December 7, 2020. We limited the search to English-language studies and, when possible, studies conducted in the United States. We restricted findings based on date of publication, with a requirement that published studies reflect work undertaken after the passing of the Olmstead ruling in 1999. The eligibility criteria were (1) mentions a person-centered non-medical measurement tool, (2) setting limited to HCBS (e.g., not medical rehabilitation or skilled nursing facilities), (3) includes samples with intellectual or developmental disability (I/DD), physical disability, psychiatric disability, age-related disability, (4) written in English, (5) peer-reviewed source, and (6) uses a United States sample. The initial search yielded 8,066 studies. After title and abstract screening, we moved 178 articles to full text review, of which we retained 25 which passed a quasi-sensitivity analysis. A fuller understanding of if and how HCBS users are living satisfying and self-determined lives requires that outcome measures be augmented to incorporate the desires, needs, and goals of users. Outcome measures should incorporate not merely user satisfaction with supports, but questions that allow for the connection between outcomes desired by people and the perception of how well supports are working to help achieved those outcomes. In other words, measures should gather information about how useful services and supports are in helping people make progress towards their desired outcomes. There are only a few efforts to do this. The University of Minnesota Research and Training Center on HCBS Outcome Measurement is developing and piloting such questions, and the Shirley Ryan AbilityLab Research and Training Center on HCBS Outcome and Measurement is developing measures of perceived effectiveness of supports. These measures will allow HCBS provider organizations to embark on and monitor quality improvement initiatives and enhance opportunities for HCBS users to drive the implementation of person-centered supports. Beyond the most immediate uses of person-centered outcomes data, there are potential implications for public reporting of data on HCBS quality, use of data for certification and accreditation purposes, as well as reporting to state and federal agencies for the purposes of value-based payment. In conclusion, while the potential for person-centered outcomes data from HCBS is obvious, better measurement instruments are needed to achieve this goal.

## Introduction

Home and community-based services (HCBS) describe a wide range of health-related and social supports and services intended to support people with disabilities and older adults to live in their own home or the community.^1^ In 2014, the Centers for Medicare and Medicaid created the HCBS Final Settings Rule, which highlights the importance of person-centered service delivery.^2^ As the demand for home and community-based services continues to grow, so too has the demand for quality measure development to evaluate HCBS. The Final Settings Rule adds another level of complexity to measurement, requiring consideration of what person-centered outcome measures could be.

In July 2022, CMS released its recommendations for HCBS Quality Measures.^3^ These recommendations represent an important step forward for improving the quality of HCBS. Nevertheless, many of the measures included in the recommended quality measure set for HCBS are not adequate measures of non-medical person-centered outcomes. Person-centered outcome measures should focus on how well a person is achieving a full and self-determined life, according to what is important *to* them. In contrast, medical outcomes usually focus on outcomes that others have identified as important *for* them. In contrast to institutional settings that often provide primarily medical services, HCBS settings, where people receive a wide variety of services and non-medical supports (e.g., personal attendant care, service coordination, job coaching, home modifications, and more) present a natural opportunity to implement person-centered measures. Policymakers, providers, stakeholders, and beneficiaries need reliable, valid, and sensitive measures to assess the quality of HCBS, and to focus on the outcomes related to a person’s desired goals. Person-centered, non-medical outcomes, that are the focus of this RRTC, can encourage provider organizations to focus on needs expressed by the beneficiary, evaluate effectiveness of services, allow participants to make educated decisions when multiple plans are available, and promote accountability. Person-centered outcomes are fluid and may have different levels of achievement at different times. Progress toward a desired outcome or goal may be as valuable as achievement. Further, as the status of these outcomes is often a subjective assessment; individuals who receive the same supports and services under the same set of circumstances may experience them quite differently, posing challenges for measuring HCBS quality.

HCBS quality measures are essential to assuring that the services provided are used in a meaningful, cost-effective manner, and that they are helping to achieve the desired outcomes. The integration of HCBS into a broader system of support for person-centered outcomes creates unique challenges for measure development. HCBS quality measures have been challenging to develop for multiple reasons. For example, the process to have quality measures approved by the National Quality Forum is complex and takes time for multiple rounds of public comment and expert review. In addition, HCBS themselves are complex, with significant variation in state requirements for who is eligible to receive HCBS, what services and settings are covered in HCBS, which providers deliver services, and service coordination processes related to planning and implementing formal and informal supports. Further, the federal requirements and guidance for person-centeredness are still evolving as compliance to the 2014 CMS HCBS Final Settings rule approaches. The Final Settings Rule will have a significant influence on final quality measure guidelines. Given the different sources of support that an individual may receive at various times, including informal supports from family, it can be difficult to determine accountability for outcomes.

Despite these challenges, several measure sets are in various stages of development and adoption. Many of those measures have addressed the achievement of outcomes, as a binary option (achieved or not). Our focus is on measuring progress toward outcomes and full achievement of those outcomes as well as measuring the effectiveness of the services received in supporting the desired outcomes.

### Background

This review was part of a larger project aimed to accelerate the development and implementation of non-medical, person-centered metrics that support quality assurance and quality improvement of Federal and State home and community-based service programs, policies, and interventions.^4^

### Objective

This review aimed to describe the extent to which HCBS outcome instruments published in the peer-reviewed literature are person-centered. We sought to identify measures that relate to progress toward or achievement of outcomes and service effectiveness. The research questions we evaluated were:

- What non-medical, person-centered outcome measures are used in HCBS?
- What non-medical, person-centered effectiveness measures of HCBS supports are used?

### Search Strategy

We worked with a research librarian to develop a bibliographic search strategy. We identified text words and controlled vocabulary terms describing person-centered measures, non-medical context, home and community-based services, HCBS instruments, as well as effectiveness and quality. We applied Boolean and proximity operators for a comprehensive, yet specific search. We modified and performed the search on Medline (Ovid); The Cochrane Library (Wiley); CINAHL Plus with Full Text (Ebsco); Scopus (Elsevier); PsycInfo (Ebsco); and ProQuest dissertations & theses global. We searched each database from date of inception to December 7, 2020. We limited the search to English-language studies and, when possible, studies conducted in the United States. We restricted findings based on date of publication, with a requirement that published studies reflect work undertaken after the passing of the Olmstead ruling in 1999. We selected this criterion to ensure that instruments were being used to evaluate services that emphasized community living, as opposed to institutional settings. We did not restrict our findings based on document type. We reviewed the reference lists of included studies for additional articles. The eligibility criteria are listed below.

### Eligibility Criteria

#### Inclusion

- Must mention a person-centered non-medical measurement tool
- Setting – Home and Community-Based Services (e.g., not medical rehabilitation or skilled nursing facilities)
- Must include a population of interest: Intellectual or Developmental Disability (I/DD), Physical Disability, Psychiatric Disability, Age-Related Disability.
- Only articles written in English were considered.
- Peer-reviewed literature
- United States population

#### Exclusion

- Non-English
- Measures intended for use only in institutional settings
- Measures that include only medical outcomes
- Abstract collections
- Dissertations
- Non-adult (younger than age 21)
- Elderly only
- Nursing homes, Skilled nursing facilities, in-patient care settings
- Palliative care
- Hospice population
- Clinical Trials
- Publication prior to Olmstead ruling (1999)

### Selection Process

We used Covidence software for the review process.^7^ Independent reviewers divided into pairs to screen the titles and abstracts for the first round. Full articles were reviewed to gather enough information if the title and abstract did not suffice when considering inclusion. The second round for eligibility included review of full text articles to assure the studies met the inclusion criteria. Covidence was used to determine agreement between the pairs. If there was not agreement, the research team met to reconcile and make final decisions about inclusion. Documentation of rationale for inclusion and exclusion of specific titles, abstracts, and full texts was performed.^5-6, 8-9^ Due to changing perceptions and understandings of person-centered outcome measures during the review process, the lead author performed a quasi-sensitivity analysis with several co-authors. This process involved: 1) reviewing articles excluded during the full-text review to confirm appropriate application of inclusion and exclusion criteria, and 2) reviewing included articles to confirm they answered the study question. Based upon this additional review, several articles that had been excluded were subsequently deemed to be appropriate for extraction, and several articles that had been included were deemed ineligible based on population of interest or year of data collection/publication.

## Findings

The initial search yielded 8,066 studies. After title and abstract screening, 178 articles moved to full-text review as the majority were irrelevant to the research question. After full-text review and the quasi-sensitivity analysis, a final sample of 25 articles was retained.

Table 1 shows the instruments that were used in the 25 articles. Some articles used a number of instruments to measure outcomes of HCBS users. The National Core Indicators (in-person survey) and the Personal Outcome Measures were used in more than one article (4 and 7 respectively). The majority of these instruments appear to have been validated for use with either the general population or with specific disability populations. The majority of instruments are administered via interview, with a few using survey questionnaires.

**Table 1.**
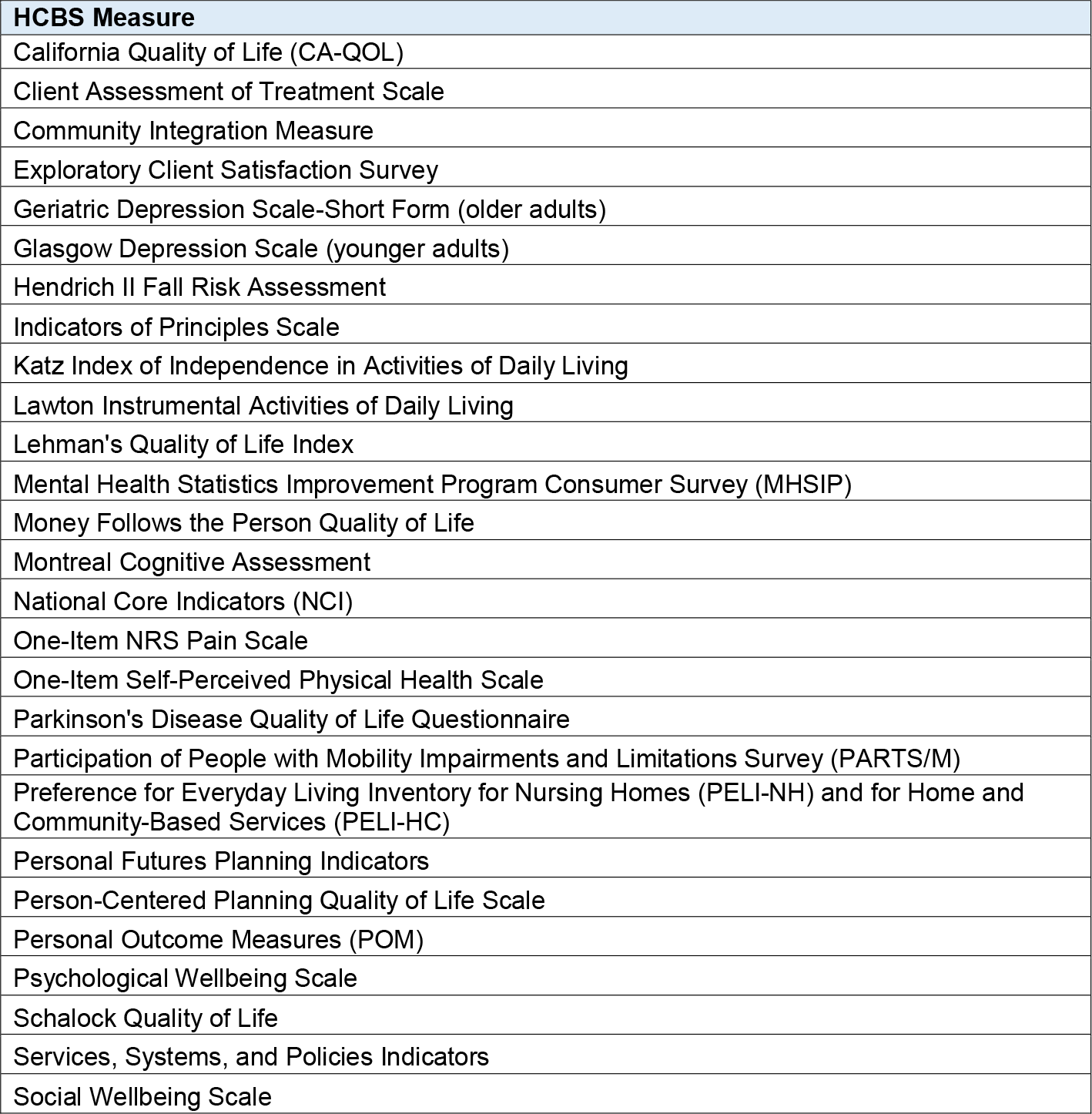
List of HCBS Measures.

Table 2 shows the populations included by the 25 articles. In some cases, the instruments capture data on people from multiple different populations, which is why the number of articles covering each population sums to more than 25. The articles most often covered people with intellectual/developmental disability, followed by people with psychiatric disability, physical disability, and age-related disability. There were two articles that did not specify the population they focused on beyond stating that it was people served on HCBS waivers.

**Table 2.** Populations included in HCBS instruments.

| HCBS population | Count of Articles that cover population |
| --- | --- |
| Age-related | 9 |
| HCBS waiver populations | 2 |
| Intellectual/Developmental Disability | 14 |
| Physical Disability | 11 |
| Psychiatric Disability | 13 |

Table 3 shows the count of instruments that included outcomes of HCBS users in the areas of community inclusion, choice and control, and holistic health and function. While some of the instruments include questions looking at different areas, we focused on identifying whether instruments addressed the outcome-oriented domains of HCBS quality from the National Quality Forum 2016 report on gaps in performance measurement.^9^ The report includes a number of domains for understanding quality of HCBS, but the domains of community inclusion, choice and control, and holistic health and functioning are the most outcome-oriented of the domains. For instruments that were included in the University of Minnesota’s RTCOM HCBS Instrument Database,^10^ we used their outcome domain classifications. For instruments not included in the database, we reviewed questions and identified which outcome most closely aligned with the question.

**Table 3.** Outcomes measured by HCBS instruments.

| HCBS Domain | Count of Instruments that cover domain |
| --- | --- |
| Community Inclusion | 14 |
| Choice and Control | 12 |
| Holistic Health and Functioning | 19 |

**Table 4.**
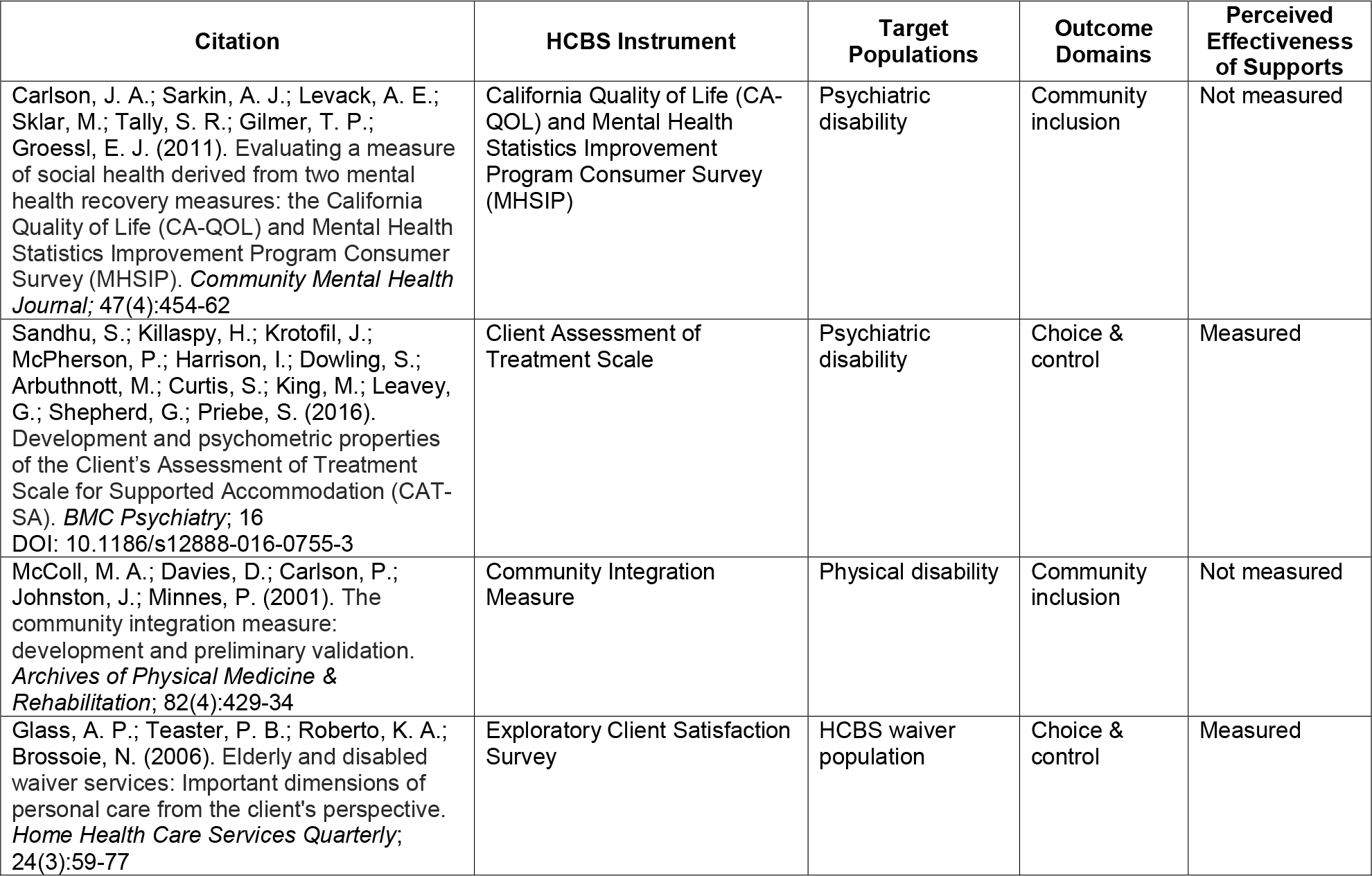
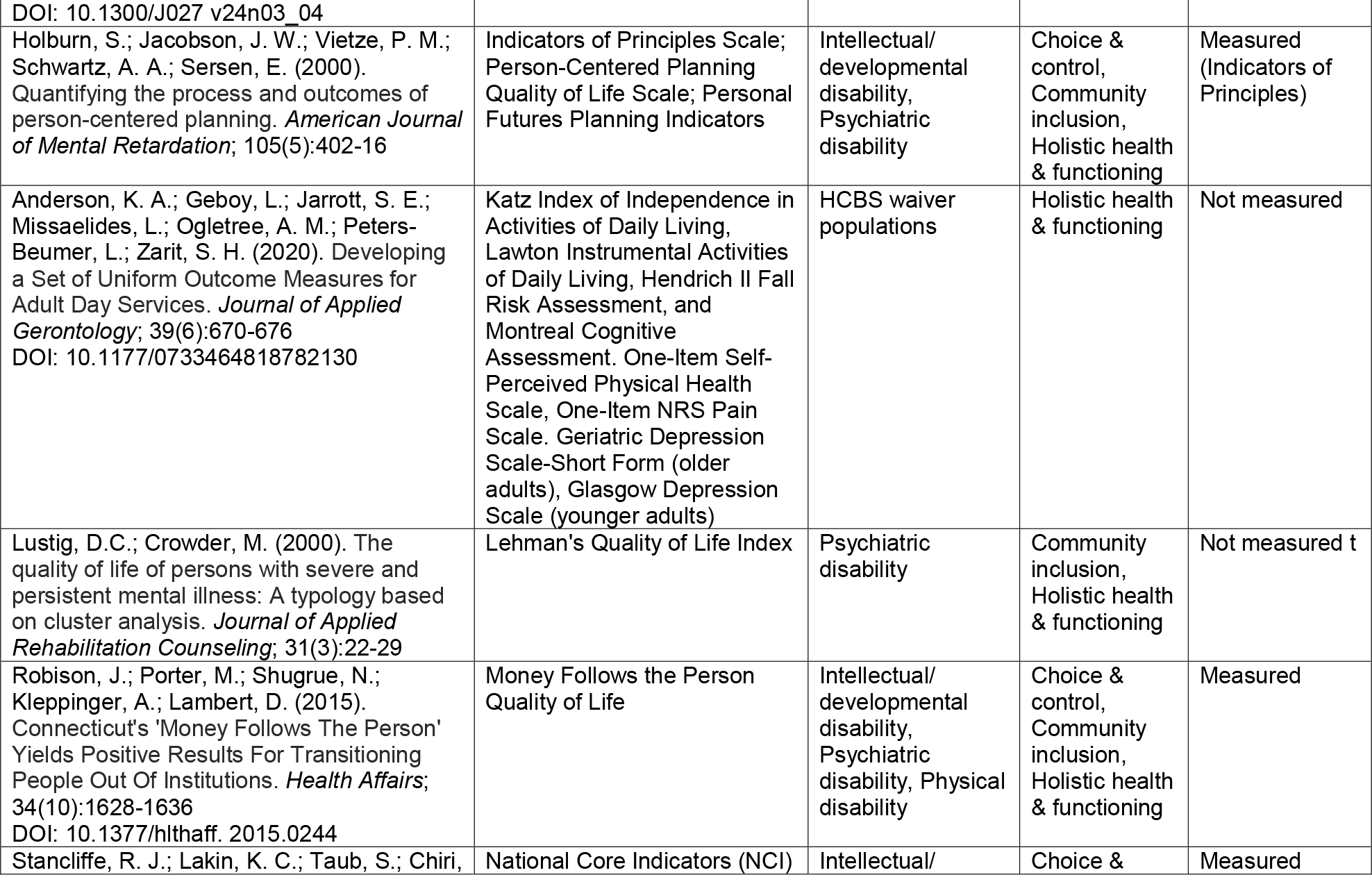
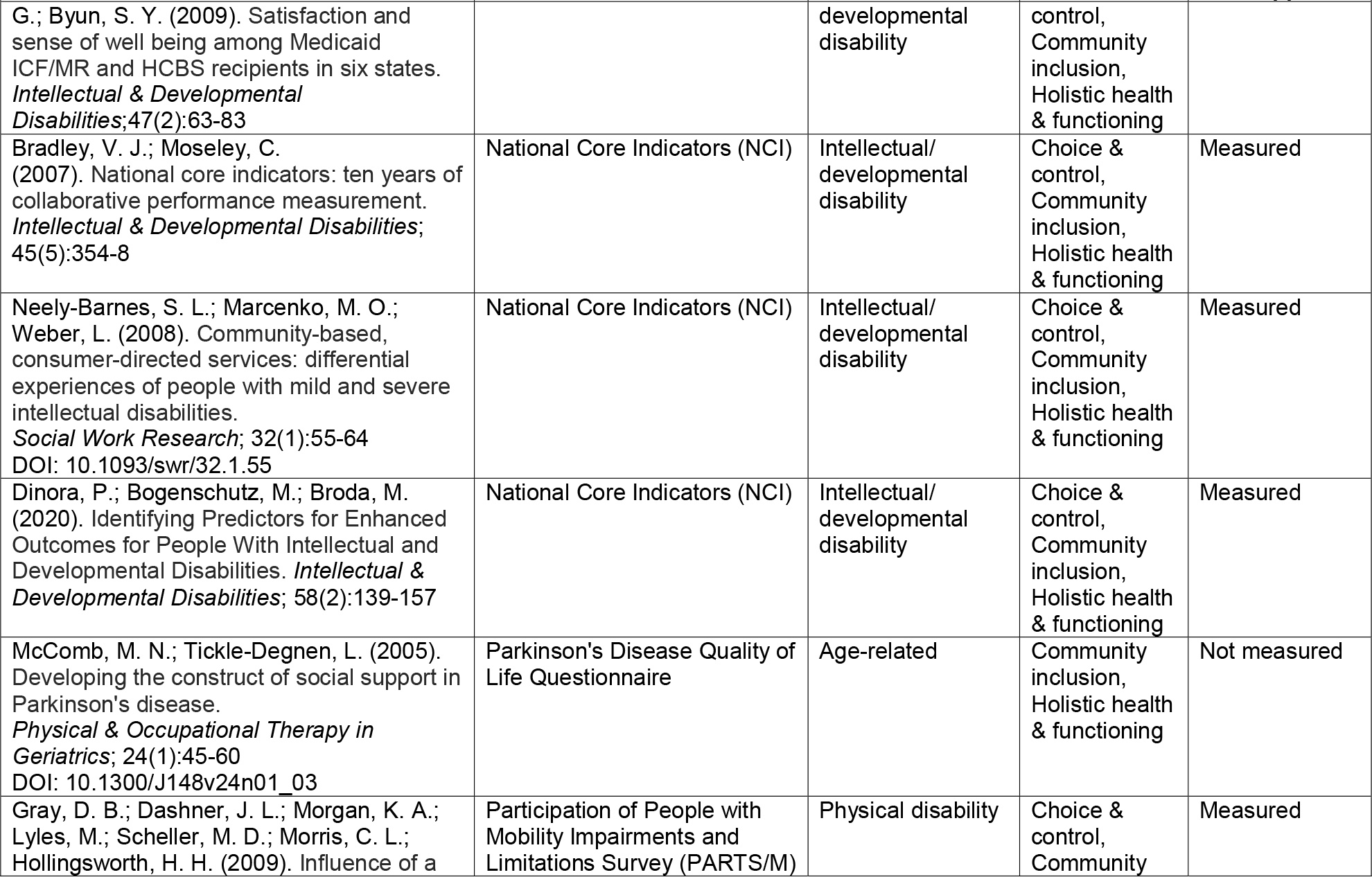
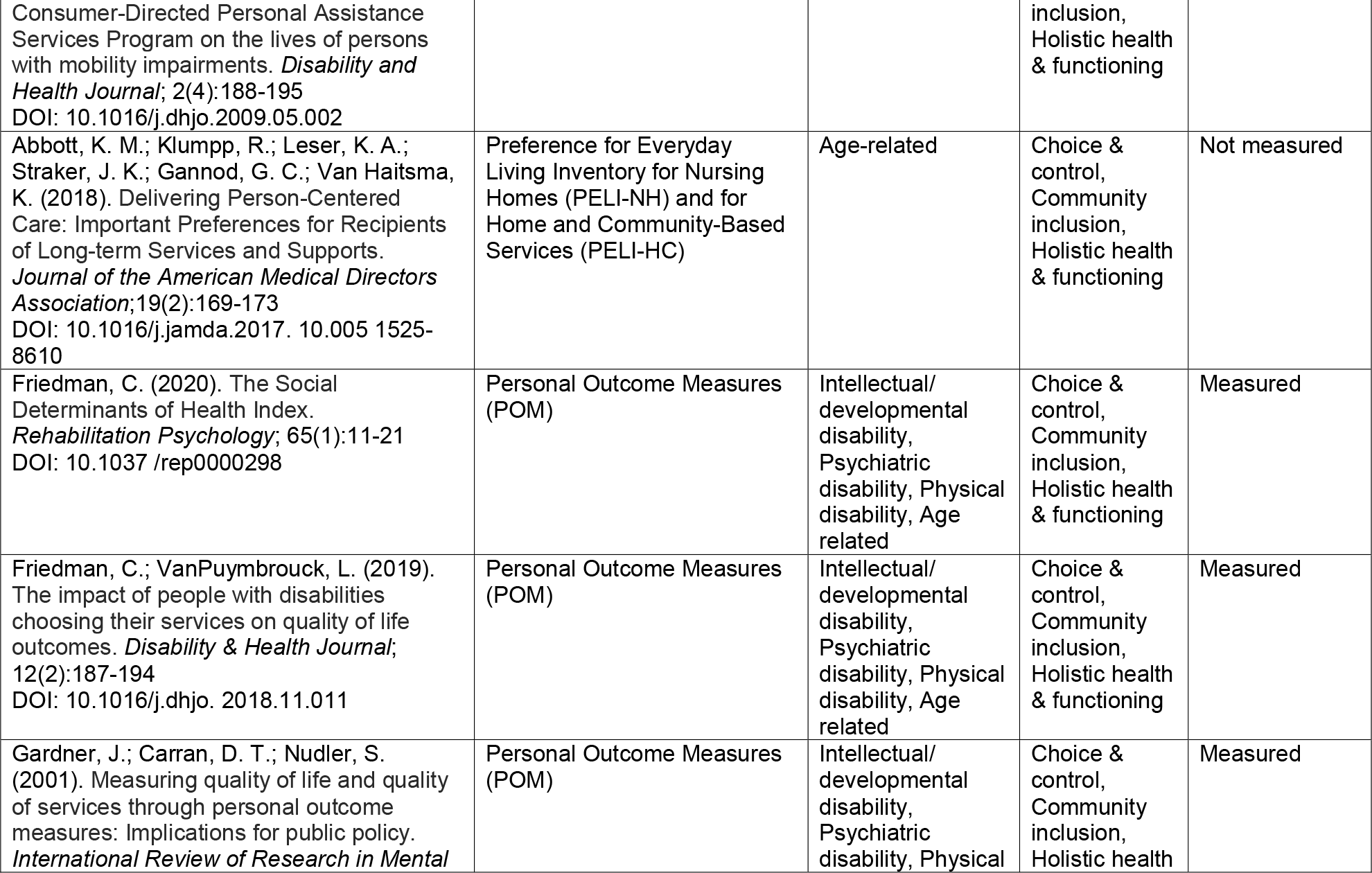
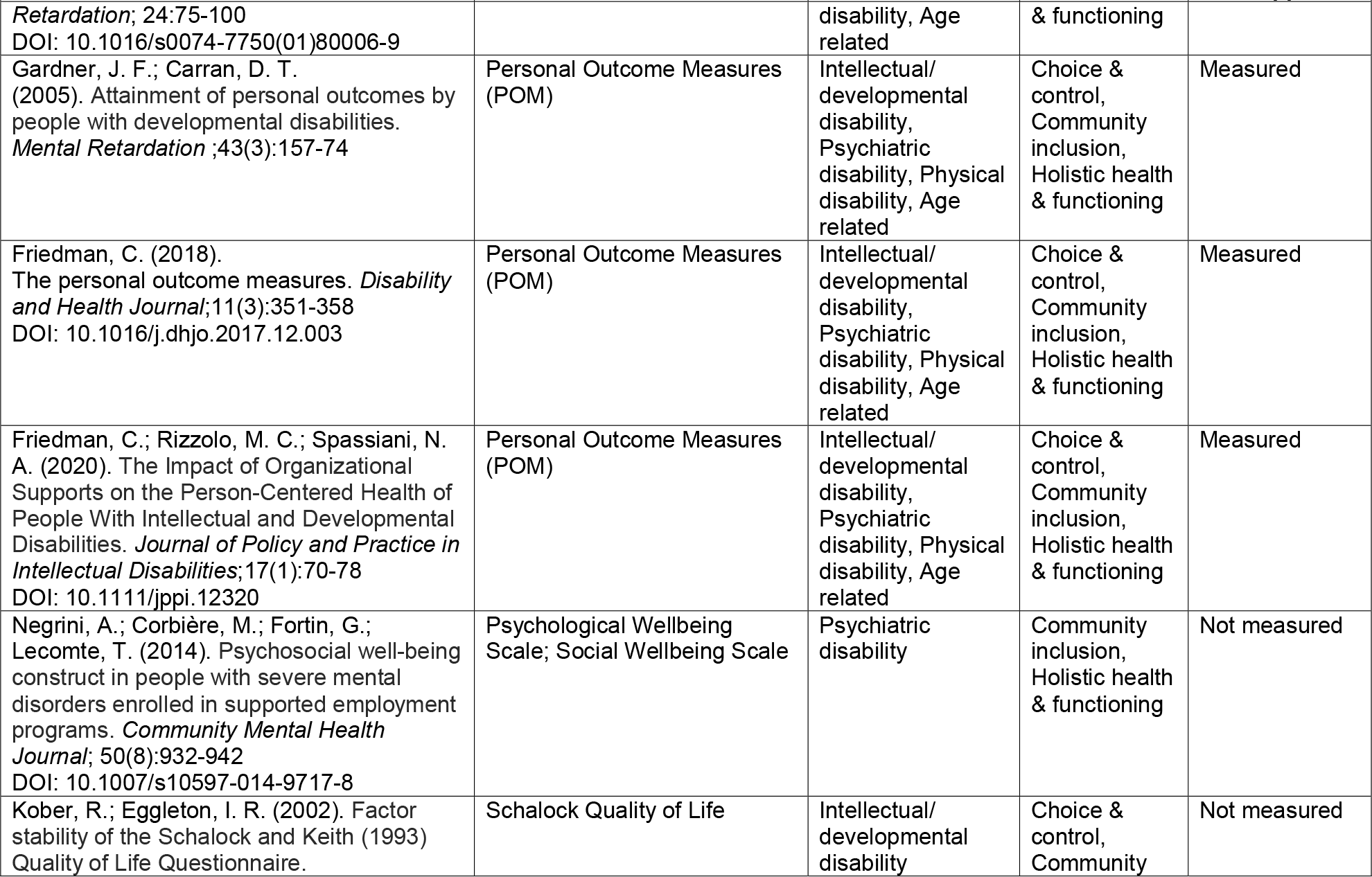
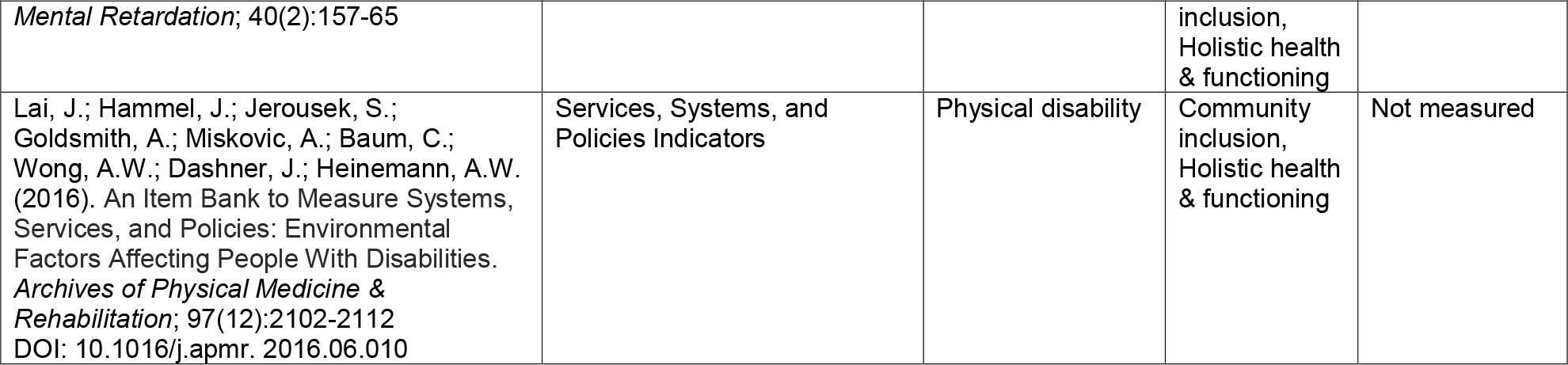
Articles Reviewed, Measurement Instruments Used, Populations Covered, Outcome Domains, and Measures of Perceived Effectiveness of Supports Included.

### Perceptions of services and supports measured by HCBS instruments

Eight instruments contained questions about the services and supports that people received. Often, these questions focused on unmet needs, desires for change, or perceptions about the quality of their providers (e.g., “Do you feel respected and regarded well?” and “Does your therapist/case manager/key-worker understand you and is he/she engaged in your treatment?”), and satisfaction with services. Of the instruments that were selected in the review, only the Personal Outcome Measures (POMs) assessed the presence of supports as they relate to key outcomes. For example, the POMs includes questions about what types of supports people have in place to achieve their desired outcomes (e.g., “What supports do you need to participate as often as you’d like in community activities?” and “What supports does the person need to develop or maintain social roles?”). The POMs also includes questions about the presence of supports as they relate to individuals’ goals, such as:

1) Does the organization know the goals the person has identified for him or herself or are efforts being made to learn about the person’s goals?
2) Does the organization provide supports and services to assist the person in pursuing personal goals?
3) Has the organization identified accomplishments the person sees as significant?
4) Does the organization assist the person to celebrate the achievement of personal milestones?

Interviewers use the “information gathering” page of the POMS manual for each indicator to guide a conversation with the person receiving supports. They also use questions from this source to guide follow-up interviews about the need and adequacy of individual supports. A decision-tree based on the person receiving supports’ information determines if the desired outcomes and needed supports are present based on the interviewees’ statements.

### Person-centeredness of HCBS outcome instruments

There are several instruments used in HCBS settings that assess specific, non-medical outcomes and perceptions about the supports delivered. While many questions included in these HCBS instruments could be considered person-centered, they are not necessarily person-driven. The person-centered questions reviewed ask HCBS users about their outcomes, but they do not identify if these outcomes were important to the user or if it was an outcome they wanted. Therefore, the outcomes measured are not person-driven. For example, asking a person “Do you like where you live?” or “Are you satisfied with the amount of time you get to spend with family and friends?” does not tell us if the person is living the life they want. They may like where they live but have a goal to live alone instead of with a roommate. They may feel good about the amount of time they spend with family and friends but still not be participating and engaging in their community in the ways that they want.

## Conclusion

A fuller understanding of if and how HCBS users are living satisfying and self-determined lives requires that outcome measures be augmented to incorporate users’ desires, needs, and goals. Outcome measures should incorporate not merely user satisfaction with supports, but questions that allow for the connection between outcomes desired by people and the perception of how well supports are working to help achieved those outcomes. In other words, measures should gather information about how useful services and supports are in helping people make progress towards their desired outcomes. There are only a few efforts to do this. The University of Minnesota Research and Training Center on HCBS Outcome Measurement is developing and piloting such questions, and the Shirley Ryan AbilityLab Research and Training Center on HCBS Outcome and Measurement is developing measures of perceived effectiveness of supports. These measures will allow HCBS provider organizations to embark on and monitor quality improvement initiatives and enhance opportunities for HCBS users to drive the implementation of person-centered supports. Beyond the most immediate uses of person-centered outcomes data, there are potential implications for public reporting of data on HCBS quality, use of data for certification and accreditation purposes, as well as reporting to state and federal agencies for the purposes of value-based payment. In conclusion, while the potential for person-centered outcomes data from HCBS is obvious, better measurement instruments are needed to achieve this goal.

## Data Availability

All data produced in the present work are contained in the manuscript

## Appendix Medline Search

1. exp *Patient-Centered Care/
2. ((person-centered or person-directed or consumer-directed or self-directed or patient-centered) adj5 (care or measure* or support* or service* or practice* or approach* or climate* or program* or policy or policies or intervention* or planning or coordination)).ti,ab.
3. 1 or 2
4. *Residential Facilities/
5. exp Assisted Living Facilities/
6. exp Group Homes/
7. exp Halfway Houses/
8. (“assisted living” or “community based” or “community engagement” or “community inclusion” or “group home*” or “halfway house*” or “home based” or “home health” or “human services system*” or legal or “long term care” or “long term services and supports” or ltss or “personal care” or “residential care” or “special care home*” or “support services” or “transportation”).ti,ab.
9. ((system or systems) adj2 “human services”).ti,ab.
10. 4 or 5 or 6 or 7 or 8 or 9
11. 3 and 10
12. exp Program Evaluation/
13. exp Quality Indicators, Health Care/
14. (“quality measure*” or “quality indicator*”).ti,ab.
15. ((program* or service* or system* or support* or workforce) adj2 (effective* or evaluat* or assess* or appropriateness or sustainability or benchmark* or quality or outcome* or metric* or accountability or performance*)).ti,ab.
16. 12 or 13 or 14 or 15
17. (“Rose-Recovery Oriented Services Evaluation” or “Agreement with Recovery Attitudes Scale” or “American Institutes for Research Self-Determination Assessment” or “Assessment of Quality of Life” or “Assistance with Caregiving” or “Attitudes Towards Treatment” or “Australian Community Participation Questionnaire” or “Burden Scale for Family Caregivers” or “CAHPS Home and Community Based Services” or “Camberwell Assessment of Need” or “Canadian Survey on Disability” or “Caregiver Strain Index” or “Caregiving Appraisal Scale” or “Cash and Counselling Demonstration Baseline Instrument” or “Caregiving in the U.S. 2015” or “Client Assessment Inventory” or “Client-Level Measures for Discretionary Programs Providing” or “Community Integration Measure” or (“Community Integration Questionnaire” and Revised) or “Community Participation Indicators” or “Community Service Attitudes Inventory” or “Consumer Recovery Outcomes System” or “Coordinated Assessment System” or “Basic Assurances” or “Personal Outcome Measures” or “Craig Handicap Assessment and Reporting Technique” or “Craig Hospital Inventory of Environmental Factors” or “Decision Making Involvement Scale” or “Disability Rating Scale” or “Empowering Arizona’s Individuals with Developmental Disabilities Consumer to Consumer Survey” or “Empowerment Affirmation Scale” or “Evaluating Social Inclusion Questionnaire” or “Experience of Care and Health Outcomes Survey” or “Family and Individual Needs for Disability Supports” or “Glasgow Outcome Scale-Extended” or “Hopkins Symptom Checklist” or “Illness Management and Recovery” or “Quality of Life Interview” or “Maryland Assessment of Recovery in People with Serious Mental Illness” or “Mayo-Portland Adaptability Inventory-4” or “Mental Health Statistics Improvement Program” or “Minnesota Department of Human Services Resident Quality of Life Interview” or “Minnesota Elderly Waiver Consumer Experience Survey” or “Minnesota Self-Determination Scales Decision-Making Preference Scale” or “Minnesota Self-Determination Scales Exercise of Control Scale” or “Minnesota Self-Determination Scales Self-Determination Environment Scale” or “MN LTSS Improvement Tool Form” or “MN Service Provider” or “MNSP-CMEvalCSSP” or “MNSP-CMEvalProviders” or “MNSP-PersonEvalCM” or “MNSP-PersonEvalCSSP” or “MNSP-PersonEvalProviders” or “Money Follows the Person” or “MOS Social Support Survey” or “National Core Indicators” or “National Health and Aging Trends Study” or “National Long Term Care Survey Community Questionnaire” or “National Study of Caregiving” or “National Survey of Older Americans Act Participants” or “New York Traumatic Brain Injury Waiver Program Participant Satisfaction Survey” or “Ohio Department of Aging Resident Satisfaction Survey” or “Older People’s Quality of Life Questionnaire” or “Oregon HCBS Individual Experience Survey” or “Participant Experience Survey Home*” or “Participant Experience Survey Mental Retardation*” or “Participant Experience Surveys Elderly and Disabled” or “Participation Assessment with Recombined Tools-Enfranchisement” or “Participation Assessment with Recombined Tools-Objective” or “Pearlin Coping Mastery Scale” or “Peer Outcomes Protocol” or “Perceived Autonomy Support” or “Perceived Competence for Recovery Scale” or “Perceived Quality of Life Scale” or “Personal Empowerment” or “Personal Experience Outcomes Integrated Interview and Evaluation System” or “PEONIES” or “Personal Family Caregiver Survey” or “Personal Life Quality Protocol” or “Personal Wellbeing Index-Intellectual Disability” or “Person-Centered Practices in Assisted Living” or “PROMIS” or (“Quality of Life” and Kane) or “Quality of Life in Alzheimer’s Disease Scale” or “Quality of Life Questionnaire” or “Quality of Life Scale” or “Recovery Assessment Scale” or “Recovery Oriented Practices Index” or “Recovery Oriented System Indicators” or “Recovery Promotion Fidelity Scale” or “Recovery Self-Assessment Scale” or “Ridgway Recovery-Enhancing Environment Measure” or “Rochester Recovery Inventory” or “Sense of Community Index 2” or “Social Acceptance Scale” or “Social and Community Opportunities Profile” or “Social Relations Scale” or “Staff Relationship Scale” or “Staff Survey of Social Inclusion” or “Stages of Recovery Instrument” or “Survey for Caregivers Supporting a Person with a Disability Outside of the Disability Support Service System” or “Tailored Caregiver Assessment” or “Temple University Community Participation Measure” or “Arc’s Self-Determination Scale” or “Empowerment Scale” or “Family Quality of Life Scale” or “Mental Health Climate Questionnaire” or “Performance Outcome Measurement Project” or “TRAIL Leisure Assessment Battery” or “UCLA Loneliness Scale” or “Well-Being Scale” or “World Health Organization Quality of Life” or “Youth Quality of Life Instrument” or “Youth Services Survey” or “Zarit Burden Interview”).ti,ab.
18. 16 and 17
19. (“Home and Community-Based Services” or HCBS).ti. 20.
20. 11 or 18 or 19
21. limit 20 to English language

## References

1. DuBois, L. & Sadler T. The Influence of Disability Models on Person-Centeredness in Home and Community-Based Services. 2022; https://www.sralab.org/sites/default/files/downloads/2022-03/HCBS%20Disability%20Models%20and%20Person-Centeredness%20Brief.pdf

2. Centers for Medicare & Medicaid Services. Federal Policy Guidance: Home and Community-Based Services Quality Measure Set. 2022. https://www.medicaid.gov/federal-policy-guidance/downloads/smd22003.pdf

3. Administration on Community Living. Grant Opportunities: https://www.grants.gov/web/grants/search-grants.html?cfda=93.433

4. Arksey H, O’Malley L. Scoping studies: towards a methodological framework. International Journal of Social Research Methodology. 2005;8(1):19–32.

5. Tricco AC, Lillie E, Zarin W, et al. PRISMA Extension for Scoping Reviews (PRISMA-ScR): Checklist and Explanation. Ann Intern Med. 2018;169(7):467–473.

6. Covidence systematic review software, Veritas Health Innovation, Melbourne, Australia. Available at http://www.covidence.org.

7. The Joanna Briggs Institute. Joanna Briggs Institute Reviewers’ Manual: 2015 edition / Supplement: Methodology for JBI Scoping Reviews. South Australia The Joanna Briggs Institute; 2015.

8. Peters MD, Godfrey CM, Khalil H, McInerney P, Parker D, Soares CB. Guidance for conducting systematic scoping reviews. Int J Evid Based Healthc. 2015;13(3):141–146.

9. Quality in Home and Community-Based Services to Support Community Living: Addressing Gaps in Performance Measurement. National Quality Forum, 2016; https://www.qualityforum.org/Publications/2016/09/Quality_in_Home_and_Community-Based_Services_to_Support_Community_LivingAddressing_Gaps_in_Performance_Mea surement.aspx

10. University of Minnesota Rehabilitation Research and Training Center on HCBS Outcome Measurement HCBS Instrument Database. https://rtcom.umn.edu/database

